# Adaptive design methods in dialysis clinical trials – a systematic review

**DOI:** 10.1101/2021.01.22.21250343

**Authors:** Conor Judge, Robert Murphy, Catriona Reddin, Sarah Cormican, Andrew Smyth, Martin O’Halloran, Martin J O’Donnell

**Author notes:** Corresponding Author Dr Conor Judge, HRB-Clinical Research Facility Galway, National University of Ireland Galway, Newcastle Road, Galway, Ireland, H91YR71.

## Abstract

**Background:** Adaptive design methods are intended to improve efficiency of clinical trials and are relevant to evaluating interventions in dialysis populations. We sought to quantify the use of adaptive designs in dialysis clinical trials.

**Methods:** We completed a full text systematic review and adhered to the Preferred Reporting Items for Systematic Review and Meta-Analysis (PRISMA) guidelines. Our review utilised a machine learning classifier and a novel full text systematic review method. We searched MEDLINE (Pubmed) and performed a detailed data extraction of trial characteristics and a completed a narrative synthesis of the data.

**Results:** 50 studies, available as 66 articles, were included after full text review. 31 studies were conducted in a dialysis population and 19 studies had renal replacement therapy as a primary or secondary outcome. While the absolute number of adaptive design methods is increasing over time, the relative use of adaptive design methods in dialysis trials is decreasing over time (6.1% in 2009 to 0.3% in 2019). Adaptive design methods impacted 52% of dialysis trials they were used in. Group sequential designs were the most common type of adaptive design method used. Acute Kidney Injury (AKI) was studied in 27 trails (54%), End Stage Kidney Disease (ESKD) was studied in 22 trials (44%) and Chronic Kidney Disease (CKD) was studied in 1 trial (2%). 26 studies (52%) were supported by public funding. 41 studies (82%) did not report their adaptive design method in the title or abstract and would not be detected by a standard systematic.

**Conclusions:** Adaptive design methods are employed in dialysis trials, but there has been a decline in their relative use over time.

**Registration Number:** PROSPERO: CRD42020163946

**Significance statement:** *What was previously known about the specific topic of the manuscript?:* The use of adaptive designs methods in dialysis trials is unquantified.

*What were the most important findings? If studies are animals, this should be specified:* Although absolute numbers of adaptive design trials have increased over time, the proportion of dialysis trials using an adaptive design has reduced. Among trials that employed an adaptive design, 52% of dialysis trials were revised due to the adaptive criteria. Group sequential designs were the most common type of adaptive design method used in dialysis randomized clinical trials. Acute Kidney Injury (AKI) was studied in 54% of trials and End Stage Kidney Disease (ESKD) was studied in 44% of trials, which used an adaptive design.

*How does the new information advance a new understanding of the kidney and its diseases?:* Adaptive design methods are effective in dialysis trials, but their relative use has declined over time.

## Introduction

### Background

Randomized clinical trials are the gold standard for evaluating efficacy, futility or harm of new therapies (1). Compared to similar specialties, nephrology has traditionally had a low number of randomized clinical trials, particularly evident for patients with End Stage Kidney Disease (ESKD) (2). The comparatively low number of trials are postulated to be due to difficult recruitment, previous history of underpowered trials and lack of funding (3,4). Although the number of trials are increasing, nephrology continues to lag behind other specialities such as cardiology, haematology/oncology and gastroenterology (5,6).

Adaptive clinical trials use interim data analyses to modify the trial design or duration in a predefined way (7), without undermining the integrity or validity of the trial, thereby preserving the Type 1 Error (False Positive) rate. The most common type of adaptive design is the Group Sequential Design (GSD), where planned interim analyses permit stopping of trials for efficacy or futility. Other designs include sample size re-estimation, multi-arm multi-stage trials, adaptive randomisation, biomarker adaptive and seamless phase II/III trials (8).

Adaptive clinical trials appear particularly suitable for the evaluation of novel interventions in ESKD, by reducing resource requirements, decreasing time to study completion and increasing the likelihood of study success i.e. power to answer hypothesis (9). Previous trials in ESKD have overly relied on observational data to inform trial design, including assumptions of expected effect size and variance (10), rather than estimates from early phase clinical trials. If incorrect, trials may be underpowered with an insufficient sample size to answer the underlying research question (10). Adaptive sample size re-estimation is a potential solution, as seen in cardiology trials (11), such as planned blinded sample size re-estimation, which identifies inaccurate assumptions, thereby triggering altered recruitment targets mid trial to ensure adequate power.

Adaptive design may also be relevant to evaluation of more established interventions. For example, the 4D trial (12) reported that atorvastatin 20mg per day did not reduce cardiovascular events in ESKD despite evidence of a 20-30% reduction in other populations (13). This trial included a single dose of statin or a single low-density lipoprotein target; it is hypothesised that alternative or multiple doses may have been more beneficial in an ESKD population given the significantly altered pharmacokinetics and pharmacodynamics (10,14). An adaptive multi-arm multi-stage (MAMS) trial design may have been more appropriate with one interim analysis at the end of stage I to identify an optimum dose to take forward into stage II. For example, the Telmisartan and Insulin Resistance in HIV (TAILoR) trial used a MAMS design with one interim analysis to identify the most appropriate dose among three telmisartan doses (20, 40 and 80mg daily). All three doses were tested in stage I and telmisartan 80mg was taken forward into stage II (15).

This systematic review aims to: (i) summarise the use of adaptive design methodology in randomized clinical trials in dialysis population and populations at risk of requiring dialysis; (ii) describe the characteristics of the adaptive designs including dialysis modality, funding, and geographical location; (iii) describe the adaptive trial design characteristics; (iv) estimate the percentage of adaptive clinical trials in dialysis among all dialysis RCTs; and (v) outline temporal trends in all the above.

## Methods

We performed a systematic review, reported according to the Preferred Reporting Items for Systematic Reviews and Meta-Analyses guidelines (PRISMA) (16). The protocol was registered with PROSPERO (CRD42020163946) and published separately (17). After testing our pre-defined search strategy (17), we found a small number (n=16) of dialysis RCTs that reported an adaptive design method. We discovered that the adaptive design methods are often not reported in the title and abstract of papers and would not be detected in a traditional systematic search. To overcome this, we developed a novel “full text systematic review” protocol and to our knowledge, this is the first use of this methodology.

### Search method for the identification of trials

#### Electronic search – Dialysis Studies

We performed an electronic search on MEDLINE (Pubmed) from database inception until 01 June 2020. Zotero was used as our reference manager. The dialysis search terms were adapted from Beaubien-Souligny et al. 2019 (18) and included dialysis, peritoneal dialysis, hemodialysis, hemodiafiltration, haemodiafiltration, hemofiltration, haemofiltration, extracorporeal blood cleansing, haemodialysis, renal dialysis, renal replacement, end stage kidney, end stage renal, stage 5 kidney and stage 5 renal (Supplementary Table 1). The output was stored in the Research Information Systems (RIS) file format.

#### Machine Learning Classifier – Randomized Clinical Trials

We used the high sensitivity machine learning classifier (RobotSearch) to identify RCTs from the dialysis search output (15). RobotSearch is a machine learning classification algorithm combining an ensemble of Support Vector Machines (SVM) and Convolutional Neural Networks (CNN) with a reported Area Under the Curve of 0.987 (95% Confidence Interval (CI), 0.984 to 0.989) for RCT classification. We adjusted the parameters of RobotSearch to perform a sensitive search to increase the proportion of RCTs that are correctly identified (15). Studies classified as likely to be randomized clinical trials were sourced for the full text systematic review.

#### Full text Systematic Review – Adaptive Design Methods

We used Recoll for Windows to perform a full text systematic review on our dialysis randomized clinical trial search results. Recoll is based on the Xapian search engine library and provides a powerful text extraction layer and a graphical interface. The adaptive design search terms were adapted from Bothwell et al., 2018 (19) and included phase ii/iii, treatment switching, biomarker adaptive, biomarker adaptive design, biomarker adjusted, adaptive hypothesis, adaptive dose-finding, pick-the-winner, drop-the-loser, sample size re-estimation, re-estimations, adaptive randomization, group sequential, adaptive seamless, adaptive design, interim monitoring, bayesian adaptive, flexible design, adaptive trial, play-the-winner, adaptive method, adaptive AND dose AND adjusting, response adaptive, adaptive allocation, adaptive signature design, treatment adaptive, covariate adaptive and sample size adjustment (Supplementary Table 2).

#### Manual full text review

We then performed manual full text review to confirm studies that were included in the final systematic review. This process is summarised in a PRISMA flowchart (Figure 1). Full text review was performed by CJ, RM and CR. Disagreements were resolved by consensus and where a resolution was not reached by discussion, a consensus was reached through a third reviewer (MOD).

**Figure 1.**
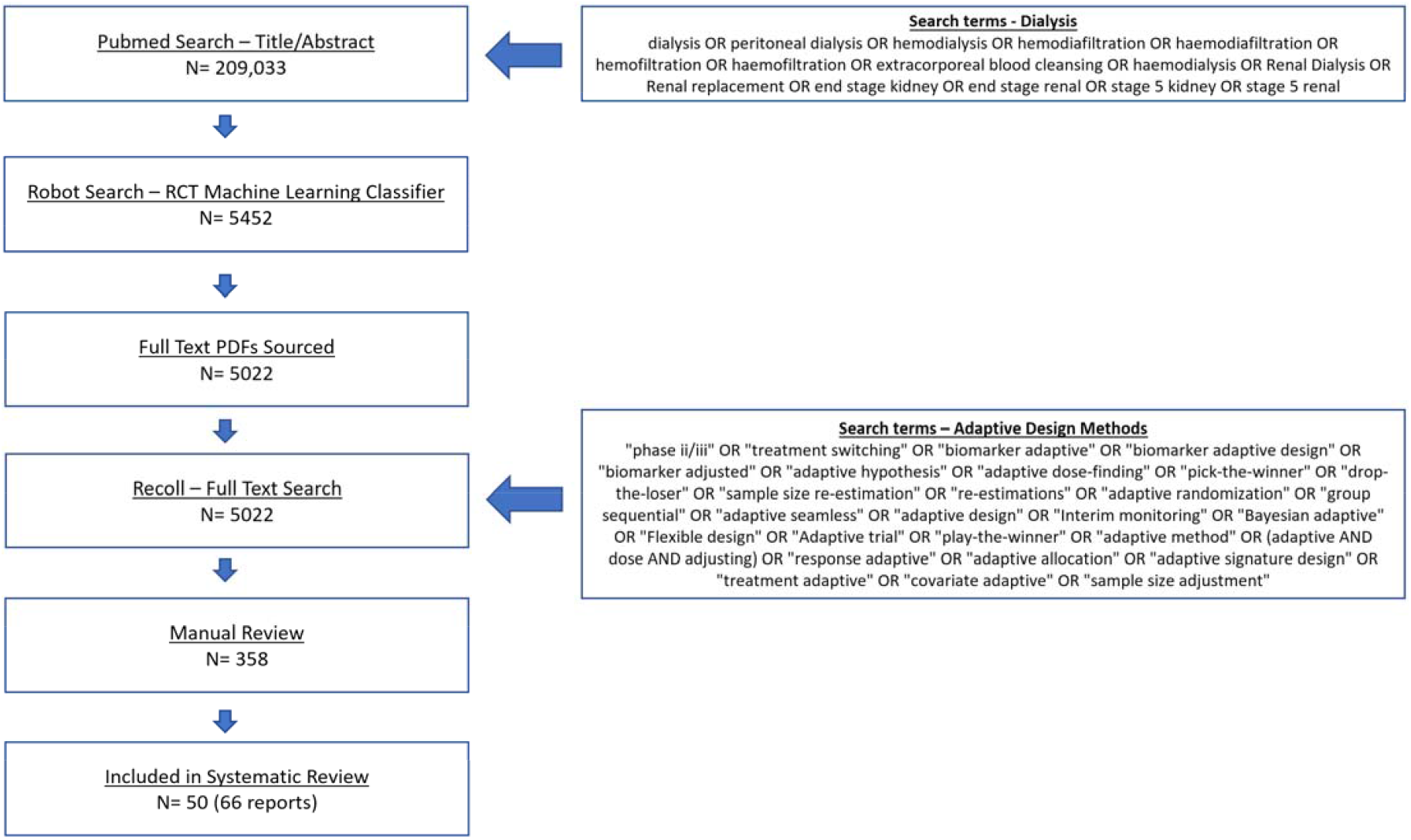
PRISMA Flow Diagram.

### Inclusion/exclusion criteria for the selection of studies

#### Type of study design and participants

Randomized clinical trials of interventions in patients with ESKD, acute kidney injury undergoing renal replacement therapy (RRT) including haemodialysis, peritoneal dialysis, haemodiafiltration and haemofiltration. We did not limit our population to any specific disease. Additionally, we included studies that included dialysis as either a primary or secondary outcome.

#### Type of intervention and outcome

We did not place a restriction on the intervention type and included trials that studied medications during dialysis, medical devices, dialysis parameters and dialysis modality. We included all outcomes including surrogate markers, patient-centred outcomes and hard clinical outcomes.

### Selection and analysis of trials

CJ, RM and CR extracted the study characteristics independently and in parallel. Data collected included the type of the adaptive design, stopping rule, impact of adaptive design (i.e., stopping for futility or efficacy, sample size changes, etc.), trial population, intervention, dialysis modality, the country of the lead investigator and the funder of the study (Adapted from Hatfield et al., 2016 (20))(Supplementary Table 3).

### Assessment of the quality of the studies: risk of bias

We used the Cochrane Risk of Bias 2 Tool (21) to assess methodological quality of eligible trials including random sequence generation, allocation concealment, blinding of participants and health care personnel, blinded outcome assessment, completeness of outcome data, evidence of selective reporting and other biases. Risk of bias assessments were performed independently by (CJ, RM, CR, SC), and disagreements were resolved by consensus. If one or more domains was rated as high, the study was considered at high risk of bias. We summarised our findings in a risk of bias table using the Revised Cochrane risk-of-bias tool for randomized trials (22)(Supplementary Table 4).

### Data synthesis

A descriptive synthesis of the data was performed. We reported overall outcomes and outcomes by (i) frequency and type of adaptive design; (ii) adaptive designs as a proportion of studies classified as dialysis RCTs by RobotSearch (iii) Population, Intervention and Outcome including dialysis modality (haemodialysis, peritoneal dialysis, haemodiafiltration and haemofiltration); (iv) publication in high impact journals; (v) geographic location and funding; (vi) reporting of adaptive design methods in title and abstract; and (vii) a risk of bias assessment.

## Results

The systematic search of articles with dialysis keywords published before 01 June 2020, identified 209,033 results. 5,452 articles were classified as probable RCTs by the machine learning classifier RobotSearch (15). Full text articles were sourced (n=5,022) and we performed a full text systematic review using adaptive design keywords which identified 358 studies for manual screening. 50 studies, available as 66 articles, were included after full text review (Figure 1). 31 studies were conducted in dialysis populations and 19 studies included RRT as a primary or secondary outcome.

### Study characteristics

#### Frequency and type of Adaptive Design

Figure 2 reports the number of adaptive design designs by year and alongside the proportion of all dialysis RCTs that used adaptive design methods. The absolute amount of dialysis trials using adaptive designs has increased each year, but this has not matched the overall increase in dialysis trials and resulted in a relative decrease over time in the use of adaptive design methods in dialysis trials ranging from 6.1% in 2009 to 0.3% in 2019 with a mean of 1.76%.

**Figure 2.**
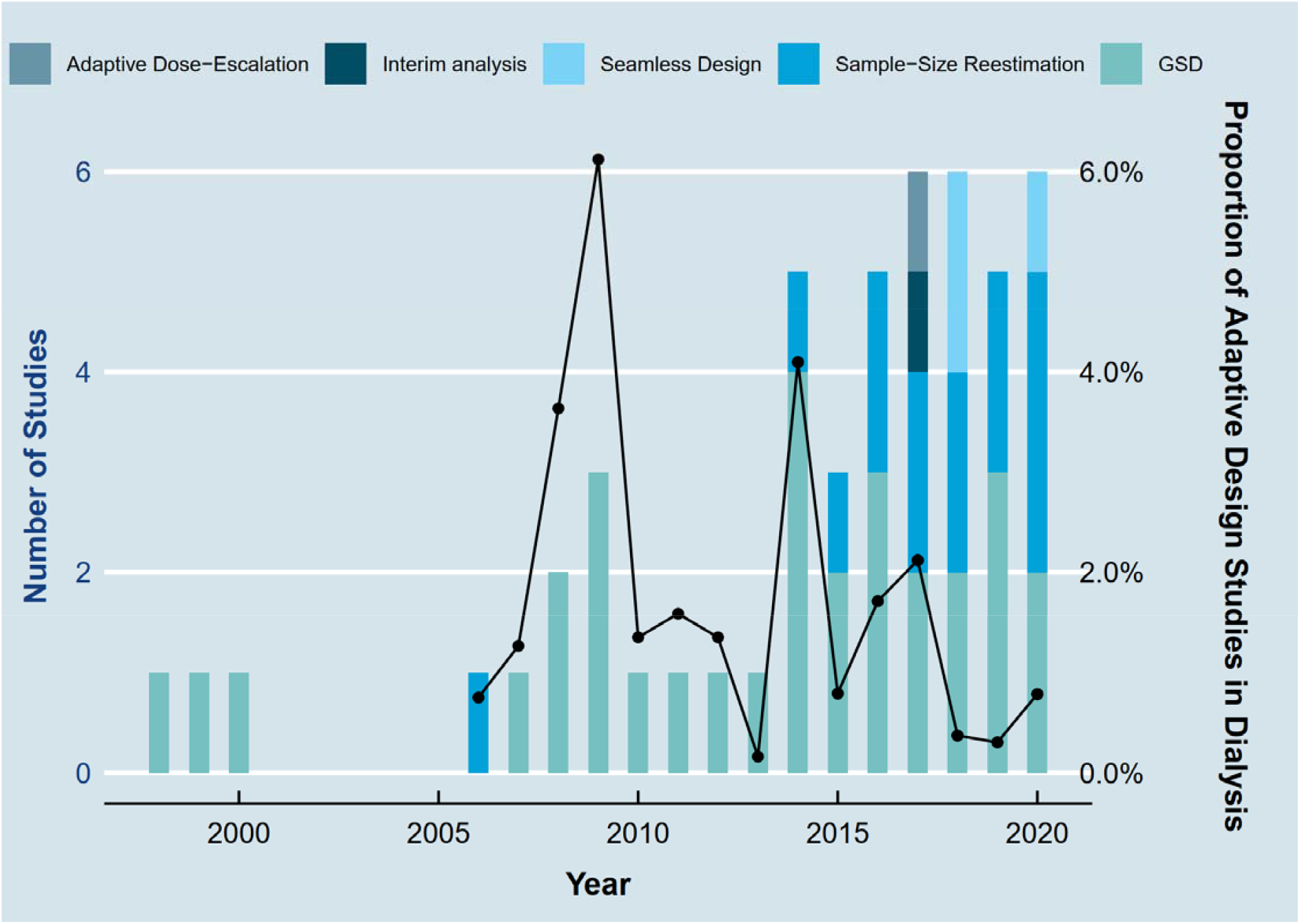
Adaptive Design in Dialysis Randomized Clinical Trials by Year.

Group sequential designs were the most common type of adaptive design method used, 31 (62%) trials (22 (71%) in dialysis populations and 9 (47.4%) in dialysis outcome trials) (Table 1). The O’Brien-Fleming stopping boundary was the most common stopping rule, used in 8 trials (25.8%), followed by Lan DeMets used in 7 trials (22.6%). 18 trials (58.1%) were impacted by the use of group sequential adaptive design including 6 trials (33.3%) that stopped early for futility, 3 trials (16.7%) that stopped early for efficacy and 4 trials (22.2%) that stopped early for safety. Sample-Size Re-estimation was the second most common type of adaptive design, used in 14 trials (28%) (8 (25.8%) in dialysis populations and 6 (31.6%) in dialysis outcome trials) (Table 2). 8 trials (57.1%) were impacted by the use of Sample-Size Re-estimation adaptive design including 5 trials (62.5%) that increased sample size. Phase II/III seamless design was the third most common type of adaptive design, 3 trials (6%) (1 (3.23%) in dialysis populations and 2 (10.52%) in dialysis outcome trials) (Table 3). Adaptive Dose-Escalation and Interim analysis were used in one trial each.

**Table 1.**
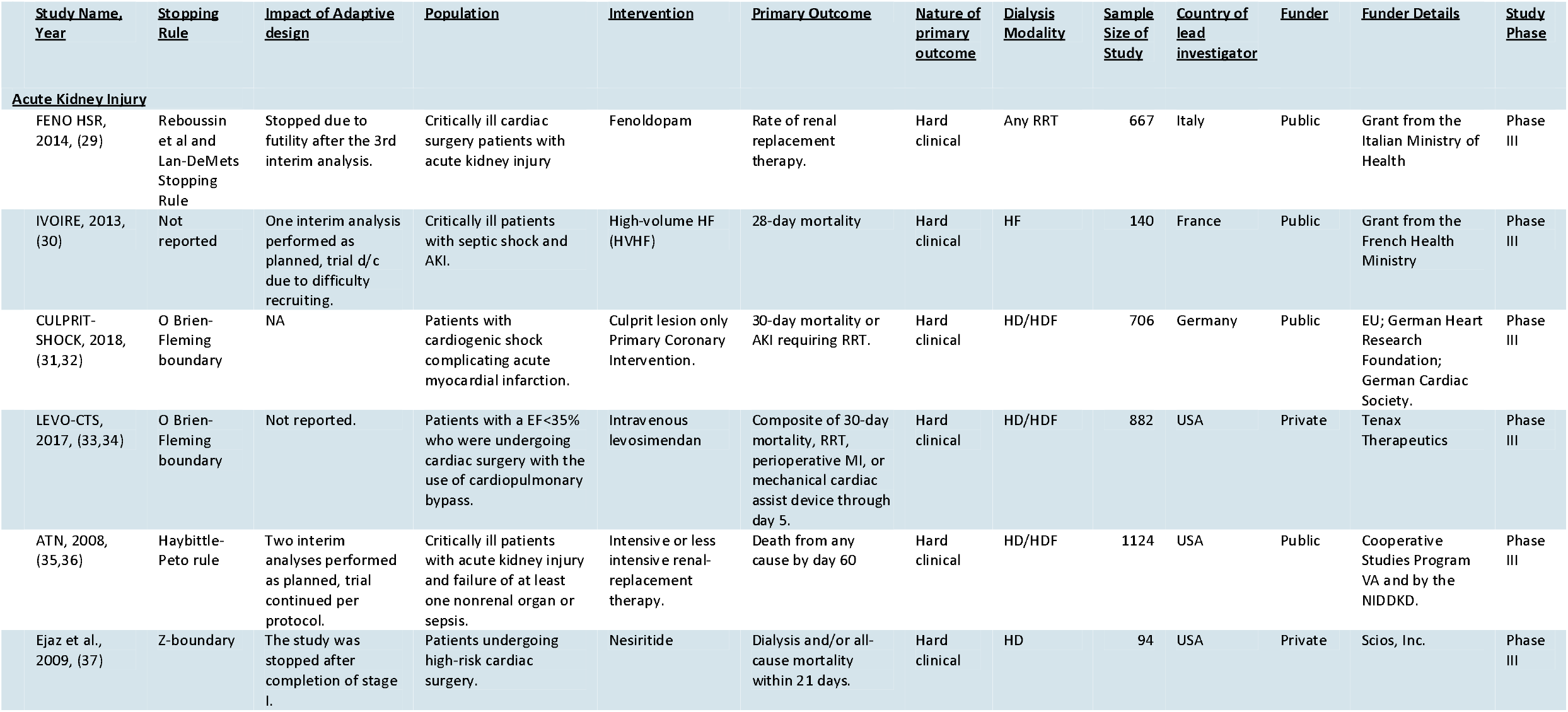
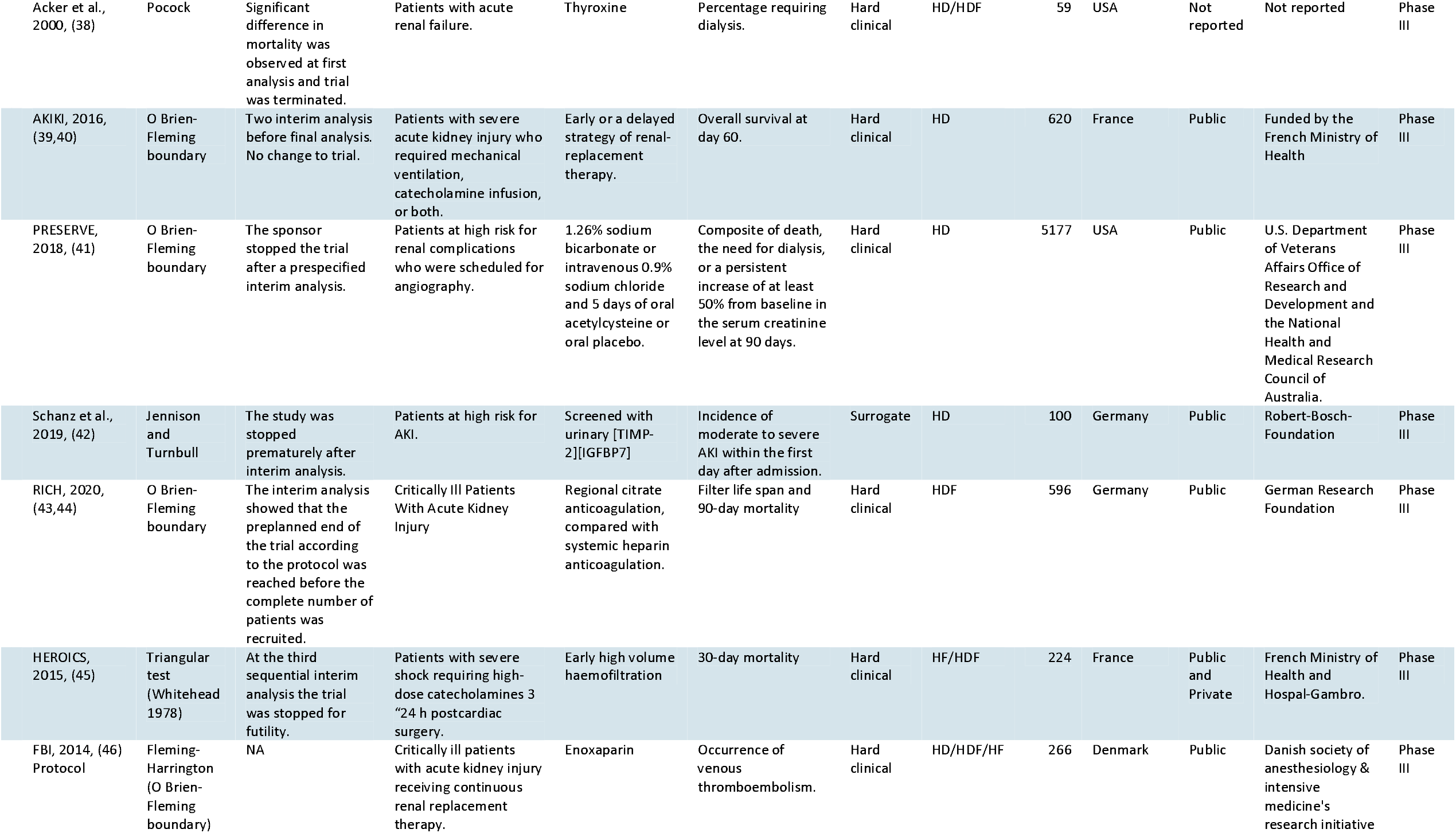
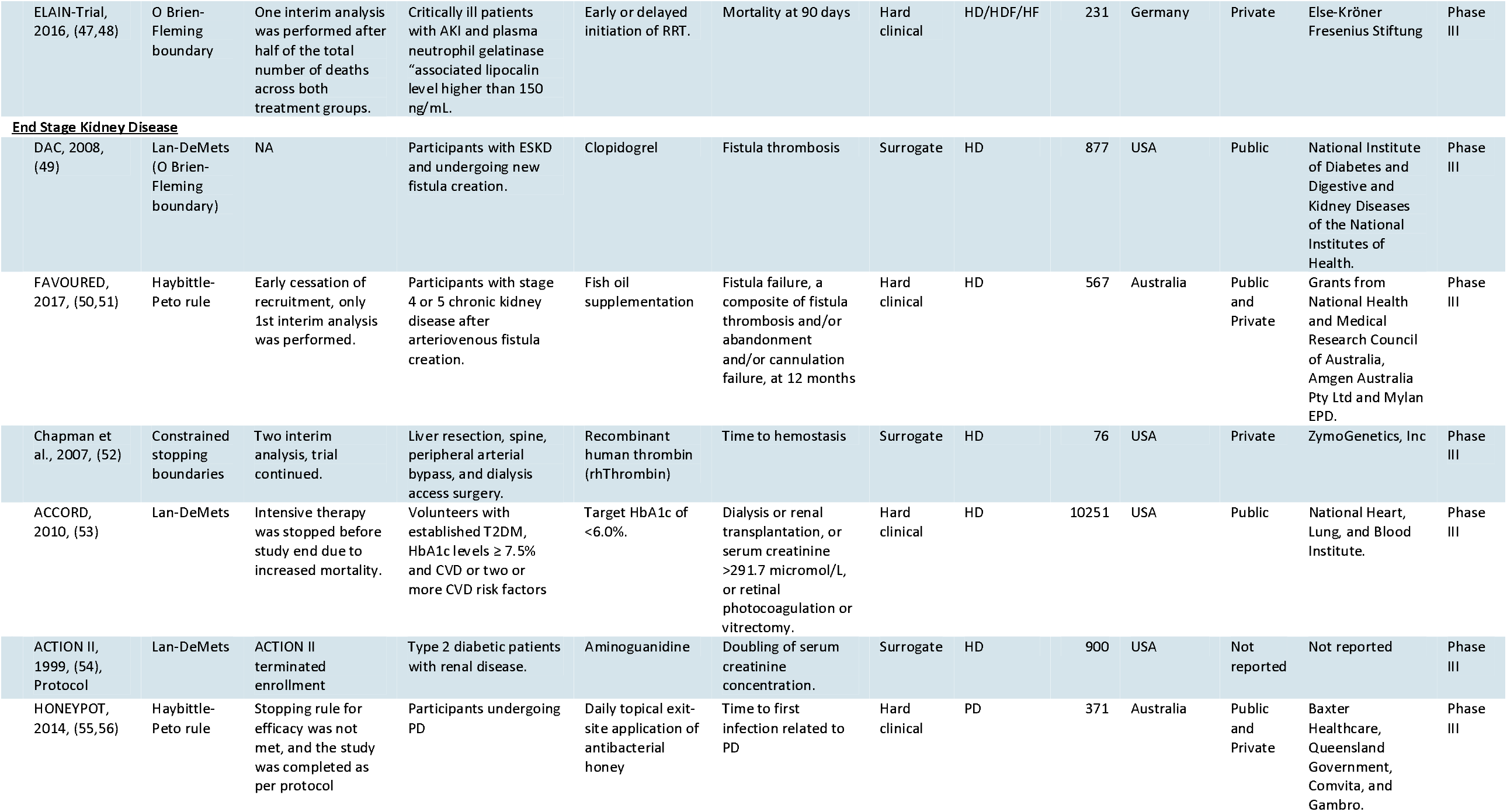
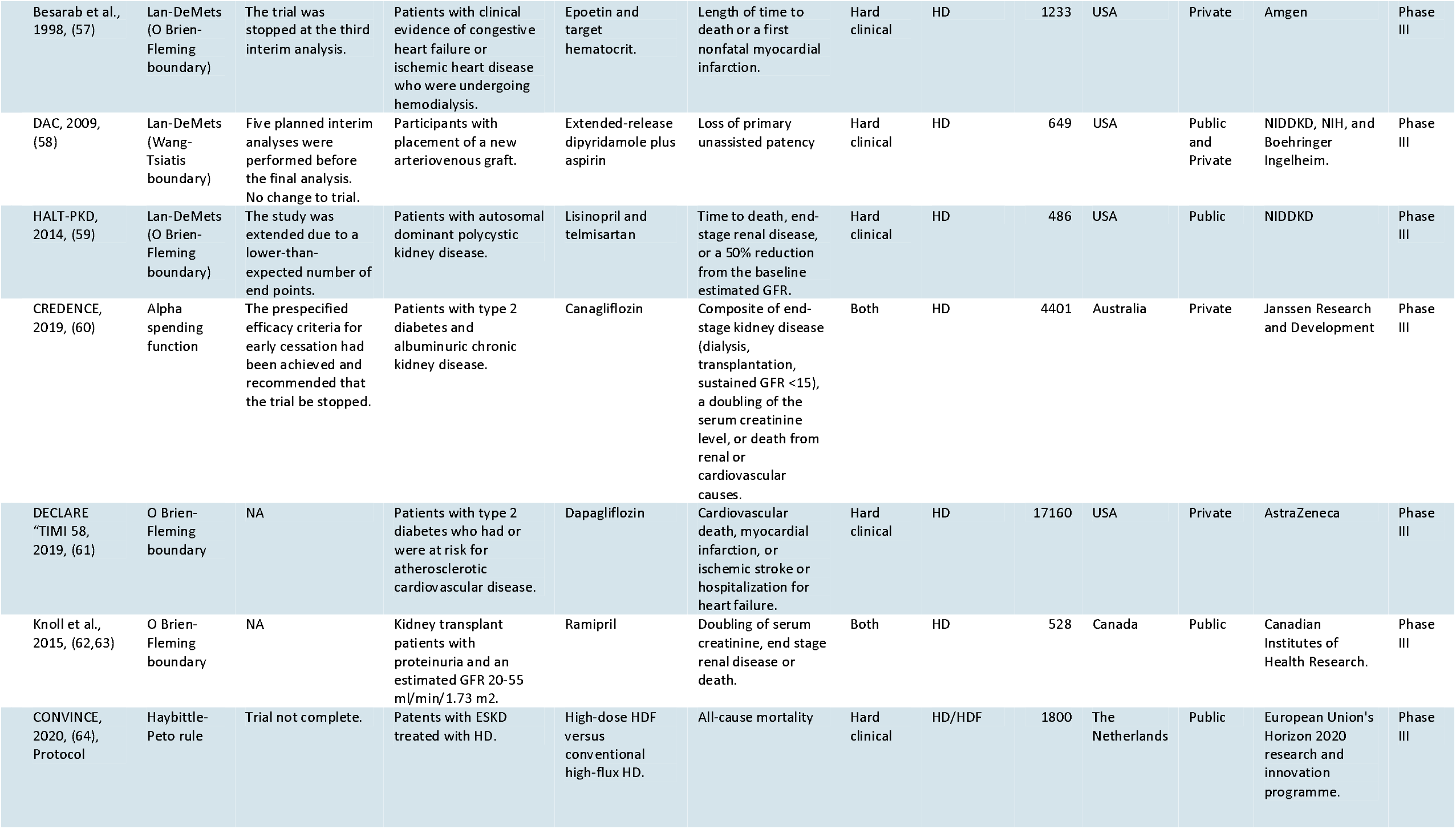
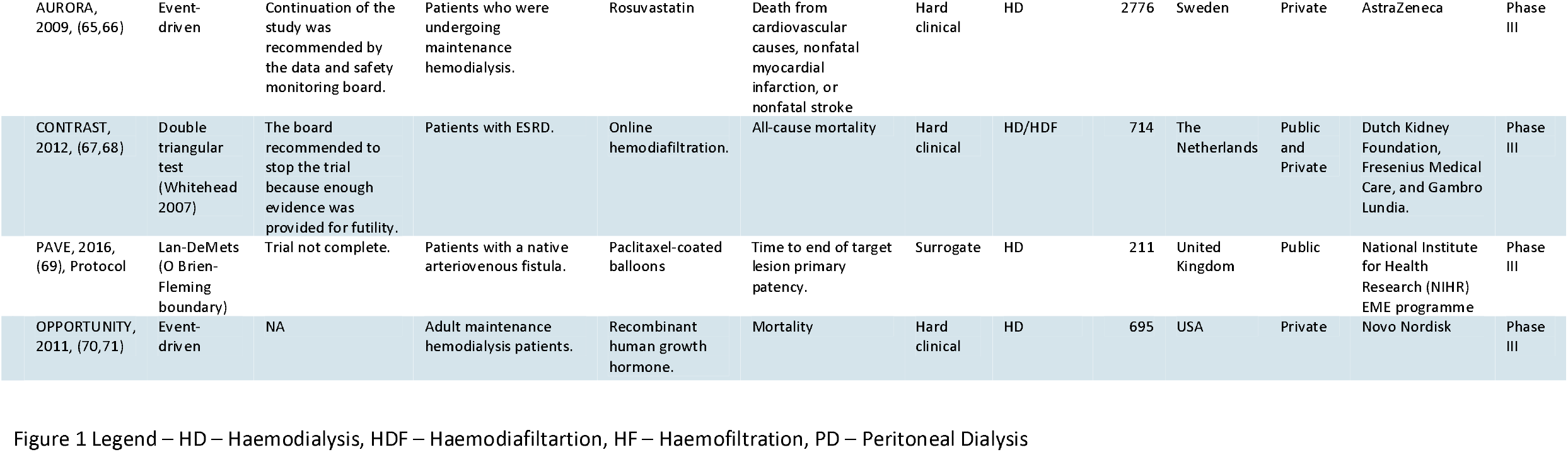
Groups Sequential Trials in Dialysis Randomized Clinical Trials

**Table 2.** Sample-Size Re-estimation in Dialysis Randomized Clinical Trials

| <u>Study Name, Year</u> | <u>Impact of Adaptive design</u> | <u>Population</u> | <u>Intervention</u> | <u>Primary Outcome</u> | <u>Nature of primary outcome</u> | <u>Dialysis Modality</u> | <u>Sample Size of Study</u> | <u>Country of lead investigator</u> | <u>Funder</u> | <u>Funder Details</u> | <u>Study Phase</u> |
| --- | --- | --- | --- | --- | --- | --- | --- | --- | --- | --- | --- |
| **Sample-Size Re-estimation** |  |  |  |  |  |  |  |  |  |  |  |
| **Acute Kidney Injury** |  |  |  |  |  |  |  |  |  |  |  |
| Kwiatkowski et al., 2017, (72) | Not reported. | Infants after congenital heart surgery. | PD | Negative Fluid Balance | Surrogate | PD | 73 | USA | Public | American Heart Association "Great Rivers Affiliate and internal funding from Cincinnati Childrens Hospital Medical Center. | Phase II |
| COACT, 2019, (73,74) | After this interim analysis, the data and safety monitoring committee advised that the sample size not be increased. | Post "cardiac arrest patients without signs of STEMI. | Immediate coronary angiography and percutaneous coronary intervention. | 90-day mortality | Hard clinical | HD/HDF | 552 | The Netherlands | Public | Netherlands Heart Institute. | Phase III |
| FRESH, 2020, (75) | NA | Patients presenting to the ED with sepsis or septic shock and anticipated ICU admission. | Dynamic assessment of fluid responsiveness (passive leg raise). | Difference in positive fluid balance at 72 hours or ICU discharge. | Surrogate | HD/HDF/HF | 124 | USA | Private | Cheetah Medical | Phase III |
| Hemodiafe, 2006, (76) | The sample size was adjusted to include 180 patients per group. | Critically ill patients with acute renal failure as part of multiple-organ dysfunction syndrome. | Intermittent HD versus continuous venovenous HDF | 60-day survival | Hard clinical | HD/HDF | 360 | France | Public | Supported by the Societe de Reanimation de Langue Francaise, with a grant from the Delegation a la Recherche Clinique de l'Assistance Publique-Hopitaux de Paris (Projet Hospitalier de Recherche Clinique) and Hospal Inc (Lyon, France). | Phase III |
| TARTARE-2S, 2016, (77), Protocol | Trial not complete. | Patients with septic shock. | Targeted tissue perfusion versus macrocirculation-guided standard care | Alive at 30 days with normal arterial blood lactate and without any inotropic or vasopressor agent. | Hard clinical | HD/HDF/HF | 200 | Switzerland | Public | Sigrid Juselius Foundation, Instrumentarium Foundation, and Helsinki University Hospital | Phase III |
| ANDROMEDA-SHOCK, 2018, (78), Protocol | Trial not complete. | Patients with septic shock. | Peripheral perfusion 'targeted resuscitation | 28 'day mortality | Hard clinical | HD/HDF/HF | 422 | Chile | Public | Departamento de Medicina Intensiva, Pontificia Universidad Católica de Chile. | Phase III |
| SCD, 2015, (79) | The study was terminated by the sponsor at the interim analysis because the SCD treatment was often outside the recommended iCa range, and therefore, resulted in ineffective therapy | ICU patients with AKI | Selective Cytopheretic Device | 60-day mortality | Hard clinical | HDF | 134 | USA | Private | CytoPherx, Inc. | Phase III |
| Riley et al., 2014, (80) | Data from the initial ten randomized patients demonstrated >50% difference in urine output, revealing adequate power would be achieved with only 20 randomized patients. | Infants <90 days old with congenital heart disease who underwent bypass surgery and were post-operatively treated with CPD. | Continue 24 h more CPD or discontinue CPD. | Urine output (ml/kg per h) | Surrogate | PD | 20 | USA | Public | Baylor College of Medicine and Cincinnati Children 's Hospital Medical Center. | Phase II |
| **End Stage Kidney Disease** |  |  |  |  |  |  |  |  |  |  |  |
| PREDICT, 2020, (81, 82) | The sample size was amended from 220 to 238 for each group. | Patients with CKD without diabetes. | High and low hemoglobin groups (Darbepoetin alfa) | Kidney composite end point (starting maintenance dialysis, kidney transplantation, eGFR<6 ml/min per 1.73 m2, and 50% reduction in eGFR). | Both | HD | 491 | Japan | Private | Kyowa Hakko Kirin, Otsuka, Dainippon Sumitomo, and Mochida. | Phase III |
| IDPN-Trial, 2017, (83) | Sample size was increased. Primary outcome was significant. | Maintenance hemodialysis patients suffering from Protein-energy wasting (PEW). | Intradialytic parenteral nutrition (IDPN) | Prealbumin | Surrogate | HD | 107 | Germany | Private | Fresenius Kabi Germany GmbH, Bad Homburg, Germany. | Phase IV |
| KALM-1, 2019, (84) | Not reported | Patients undergoing hemodialysis who had moderate-to-severe pruritus. | Intravenous difelikefalin | 24-hour Worst Itching Intensity Numerical Rating Scale (WI-NRS) | Surrogate | HD | 378 | USA | Private | Cara Therapeutics | Phase III |
| CHART, 2018, (85,86) | NA | Urologic patients undergoing elective cystectomy. | Albumin 5% or balanced hydroxyethyl starch 6% | Ratio of serum cystatin C between the last visit at day 90 and the first preoperative visit. | Surrogate | HD | 100 | Germany | Private | CSL Behring GmbH | Phase III |
| Kratochwill et al., 2016, (87) | Led to premature termination of patient recruitment | Stable PD outpatients. | Alanyl-glutamine addition to glucose-based PD Fluid. | Heat-shock protein 72 expression | Surrogate | PD | 20 | Austria | Public | ZIT - Technology Agency of the City of Vienna and FFG - the Austrian Research Promotion Agency | Phase II |
| Fujimoto et al., 2020, (88) | The sample size was calculated by the intermediate analysis of the first 30 samples enrolled. | Patients undergoing maintenance hemodialysis thrice/week. | Lidocaine/prilocaine cream (EMLA). | Puncture pain relief, which was measured using a 100-mm visual analog scale. | Surrogate | HD | 66 | Taiwan | Public | Grant-in-Aid for Young Scientists from the Japan Society for the Promotion of Science. | Phase II |

**Table 3.** Seamless Design/Adaptive Dose-Escalation in Dialysis Randomized Clinical Trials

| <u>Study Name,</u><br><u>Year</u> | <u>Impact of Adaptive</u><br><u>design</u> | <u>Population</u> | <u>Intervention</u> | <u>Primary Outcome</u> | <u>Nature of</u><br><u>primary</u><br><u>outcome</u> | <u>Dialysis</u><br><u>Modality</u> | <u>Sample</u><br><u>Size of</u><br><u>Study</u> | <u>Country of</u><br><u>lead</u><br><u>investigator</u> | <u>Funder</u> | <u>Funder Details</u> | <u>Study</u><br><u>Phase</u> |
| --- | --- | --- | --- | --- | --- | --- | --- | --- | --- | --- | --- |
| <u>Seamless Design/Adaptive Dose-Escalation</u> |  |  |  |  |  |  |  |  |  |  |  |
| <u>Acute Kidney Injury</u> |  |  |  |  |  |  |  |  |  |  |  |
| <u>Two-stage seamless adaptive design</u> |  |  |  |  |  |  |  |  |  |  |  |
| Himmelfarb et al., 2018, (89) | At the end of each stage, data from the patients are used to select the THR-184 dose arms for next stage. | Patients at high risk for AKI after cardiac surgery. | THR-184 | Proportion of patients who developed AKI | Hard clinical | HD/HDF/HF | 452 | USA | Private | Thrasos Therapeutics, Inc. | Phase II |
| <u>Interim analysis</u> |  |  |  |  |  |  |  |  |  |  |  |
| Hosgood et al., 2017, (90), Protocol | Trial not complete. | Patients receiving a kidney from a donation after circulatory death donor. | Ex vivo normothermic perfusion | Rates of delayed graft function (DGF) defined as the need for dialysis in the first week post-transplant. | Surrogate | HD | 400 | United Kingdom | Public | Kidney Research UK; University of Cambridge and University Hospitals of Cambridge Foundation Trust. | Phase II |
| <u>Phase II/III seamless design</u> |  |  |  |  |  |  |  |  |  |  |  |
| COMBAT-SHINE, 2020, (91), Protocol | Trial not complete. | Patients with septic shock-induced endotheliopathy. | Infusion of iloprost | Mean daily modified Sequential Organ Failure Assessment (SOFA) score | Surrogate | HD | 384 | Denmark | Public | Danish Independent Research Organisation | Phase II |
| <u>Phase IIa/IIb seamless design</u> |  |  |  |  |  |  |  |  |  |  |  |
| STOP-AKI, 2018, (92,93) | NA | Critically ill patients with sepsis associated AKI. | Human recombinant alkaline phosphatase | Area under the time-corrected endogenous creatinine clearance curve from days 1 to 7. | Surrogate | HD | 301 | The Netherlands | Private | AM-Pharma | Phase IIa/IIb |
| <u>Adaptive Dose-Escalation</u> |  |  |  |  |  |  |  |  |  |  |  |
| EMPIRIKAL, 2017, (94), Protocol | Trial not complete. | Patients after receiving cadaveric renal allografts. | Mirococept | Delayed graft function | Surrogate | HD/HDF/HF | 560 | United Kingdom | Public | Medical Research Council | Phase II |

#### Population, Intervention, and Outcome studied

Acute Kidney Injury (AKI) was studied in 27 trials (54%), End Stage Kidney Disease (ESKD) was studied in 22 trials (44%) and Chronic Kidney Disease (CKD) was studied in 1 trial (2%). Figure 3 reports the number of each population under study per year and shows a larger increase in adaptive design methods in AKI populations compared to ESKD populations. Medications were the most common intervention type, evaluated in 28 trials (56%), followed by Dialysis Modality in 6 trials (12%), and Dialysis Parameter in 4 trials (8%). Haemodialysis was the most common dialysis modality studied in 26 trials (52%), followed by Haemodialysis and haemodiafiltration in 8 trials (16%), Haemodialysis, haemodiafiltration and haemofiltration in 7 trials (14%) and Peritoneal Dialysis in 4 trials (8%). Hard clinical outcomes were selected in 30 trials (60%), followed by surrogate outcomes in 17 trials (34%) and mixed in 3 trials (6%). The outcome measure was continuous in 11 trials (22%) and dichotomous in 39 trials (78%). Phase III studies were the most common study phase, studied in 40 trials (80%) (Table 1-3).

**Figure 3.**
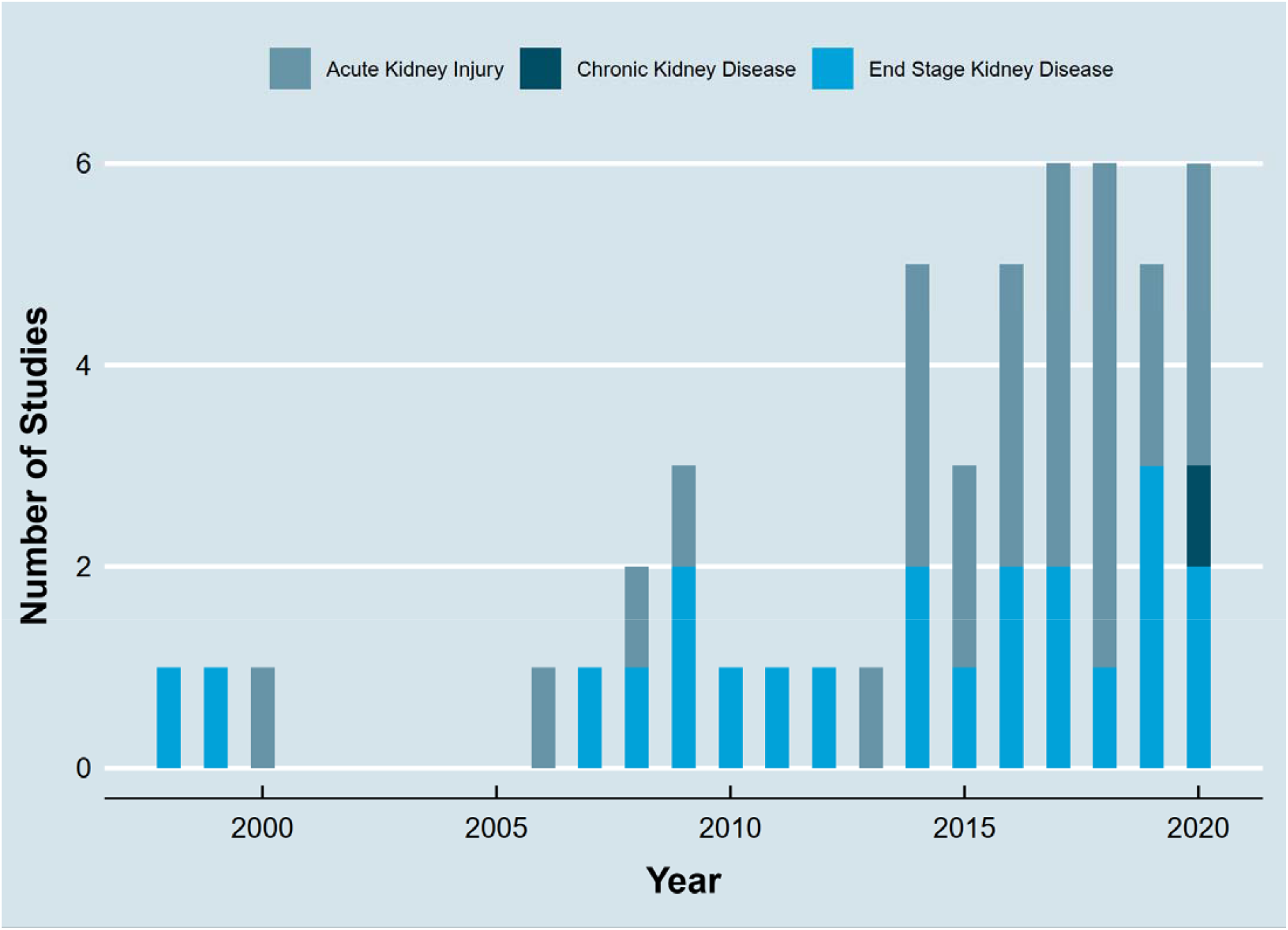
Populations with Adaptive Design in Dialysis Randomized Clinical Trials by Year.

#### Publication in High Impact Journals

30 studies (60%) were published in a high impact journal (Impact Factor > 9). 13 studies (26%) were published in the New England Journal of Medicine (NEJM), 5 studies (10%) were published in the Journal of the American Medical Association (JAMA), 4 studies (8%) were published in Trials, and 2 studies (4%) were published in the Journal of the American Society of Nephrology (JASN).

#### Geographic location and Funding

The most common country of the lead author was the United States of America in 20 studies (40%), followed by Germany in 6 studies (12%), France in 4 studies (8%), The Netherlands in 4 studies (8%), Australia in 3 studies (6%), and the United Kingdom in 3 studies (6%) (Tables 1-3). 43 studies (86%) were multicentre trials. 26 studies (52%) were supported by public funding, 17 studies (34%) were supported by private funding, 5 studies (10%) were supported by both public and private funding and 2 studies (4%) did not report the source of funding.

### Reporting of Adaptive Design Method in Title and Abstract

41 studies (82%) did not report their adaptive design method in the title or abstract and would not be detected by a standard systematic review search.

### Risk of Bias

Risk of bias was assessed for forty trials (protocols were excluded) (Supplementary Figure 1, Supplementary Table 4). Overall risk of bias was deemed to be “low” in 17 trials (42.5%), “some concerns” in 13 trials (32.5%), and “high risk” in 10 trials (25%). The randomization process led to some concerns for 10 studies (25%). Deviations from intended interventions led to some concerns for 4 studies (10%) and “high risk” for 6 studies (15%). Missing outcome data were deemed to be “some concerns” for 2 studies (5%) trials and ‘high risk’ of bias for 2 studies (5%). Measurement of outcome measures were deemed to be “some concerns” for 2 studies (5%) trials and ‘high risk’ of bias for 1 study (2.5%). Selection of the reported result were deemed to be “some concerns” for 6 studies (15%) trials and ‘high risk’ of bias for 1 study (2.5%).

## Discussion

In this systematic review, we report that adaptive design methods were used in 50 dialysis randomized clinical trials over a 20-year period. While the absolute number has increased over time, the relative use of adaptive design methods in dialysis RCTs has decreased.

First, we report that the relative proportion of adaptive design methods in dialysis trials has reduced over time. The absolute amount of dialysis trials using adaptive designs has increased each year, but this has not matched the overall increase in dialysis trials and therefore resulted in a relative decrease. We were unable to compare this result with other specialities because recent systematic reviews have not reported the relative use of adaptive designs (20,23). Second, we report that group sequential designs are the most used type of adaptive design in dialysis trials. This is similar to previous systematic reviews in cardiology (24), oncology (23) and in a review of registered clinical trials covering multiple specialities on clinicaltrials.gov (20). Third, we report that adaptive designs were more common in Acute Kidney Injury (AKI) (54% of trials) than End Stage Kidney Disease (ESKD) (44% of trials). This may reflect increasing use of adaptive design methodology in critical care (25) and sepsis related trials (26), where AKI is most common. There were very few trials of CKD with a dialysis outcome (2%) that used an adaptive design. Many reasons for the paucity of CKD trials have been previously suggested including the use of treatments in CKD despite a lack of evidence, difficulty recruiting to CKD trials due to stringent eligibility criteria and underpowered subgroup analysis (3,27). The infrequent usage of adaptive designs in CKD trials may become a self-perpetuating barrier to using adaptive designs in future trials (20). Fourth, we report that adaptive design methods impacted the conduct of the randomised trial in the majority of studies (52%). For example, 18 (58.1%) trials were impacted by the use of group sequential adaptive design including 6 trials (33.3%) stopped early for futility, 3 trials (16.7%) stopped early for efficacy and 4 trial (22.2%) stopped early for safety. This finding is similar to a systematic review of published and publicly available trials where the most common reason for stopping group sequential trials was futility (19). Fifth, we found that most common country of the lead author was the United States of America, 20 studies (40%) and the most common funding source was public, 26 studies (52%). This finding was different to a systematic review of published and publicly available trials where 65% of trials reported industry funding (19). Funding for kidney research reached an all-time low in 2013 (4), but this has recently changed in the United States with advocacy from scientific societies such as the American Society of Nephrology, an executive order to reform the US End-Stage Kidney Disease treatment industry was signed in 2020 (28). Adaptive designs are one part of the solution for optimizing the design of clinical trials in dialysis and nephrology and will benefit from the improvement in the funding landscape (27).

### Limitations

Our study has several limitations. First, we limited our search to one database (Pubmed) due to the scale of studies sourced (209,033 results). This was a deviation from our protocol but necessary to make this full text review feasible. Second, we decided to include randomised clinical trials with RRT outcomes in addition to patients currently on dialysis. This permitted a more comprehensive review of the full landscape of AKI, ESKD and CKD trials, but was a deviation from our original protocol. Third, the denominator for calculating the proportion of adaptive designs in all dialysis RCTs, will include some false negatives i.e., either not RCTs or nor dialysis. We modified the parameters of the machine learning classifier to perform a sensitive search to include as many true positives as possible. We expect this mis-classification bias to be independent of time and bias every year equally and therefore not affect the trend.

### Strengths

We developed a novel full text systematic review search strategy. 41 studies (82%) did not report their adaptive design method in the title or abstract and would not be detected by a standard systematic review search methodology. This could introduce a pseudo reporting bias where adaptive design methods are reported in the main paper but not in the abstract. Our novel strategy combined classical systematic review, machine learning classifiers and a novel full text systematic review. This new method has broad applications in medical evidence synthesis and evidence synthesis in general.

## Conclusion

Adaptive design methods improve efficiency of randomised clinical trials in dialysis, but their relative use is decreasing over time. Greater knowledge of adaptive design examples in dialysis will further improve uptake in dialysis randomised clinical trials.

## Data Availability

Extracted data is available on request.

## Author contributions

CJ, RM and MOD designed the study. CJ drafted the manuscript. CJ, RM, CR, SC, AS, MOH, MOD reviewed and approved the final version of the manuscript.

## Acknowledgments

None

## References

1. Bothwell LE, Greene JA, Podolsky SH, Jones DS. Assessing the Gold Standard — Lessons from the History of RCTs. N Engl J Med. 2016 Jun 2;374(22):2175–81.

2. Kovesdy CP. Clinical trials in end-stage renal disease—priorities and challenges. Nephrol Dial Transplant. 2019 Jul 1;34(7):1084–9.

3. Baigent C, Herrington WG, Coresh J, Landray MJ, Levin A, Perkovic V, et al. Challenges in conducting clinical trials in nephrology: conclusions from a Kidney Disease—Improving Global Outcomes (KDIGO) Controversies Conference. Kidney Int. 2017 Aug 1;92(2):297–305.

4. Bryan L, Ibrahim T, Zent R, Fischer MJ. The Kidney Research Predicament. J Am Soc Nephrol JASN. 2014 May;25(5):898–903.

5. Chatzimanouil MKT, Wilkens L, Anders H-J. Quantity and Reporting Quality of Kidney Research. J Am Soc Nephrol. 2019 Jan 1;30(1):13–22.

6. Yaseen M, Hassan W, Awad R, Ashqar B, Neyra J, Heister T, et al. Impact of Recent Clinical Trials on Nephrology Practice: Are We in a Stagnant Era? Kidney Dis. 2019;5(2):69–80.

7. Chow S-C, Chang M, Pong A. Statistical Consideration of Adaptive Methods in Clinical Development. J Biopharm Stat. 2005 Jul 1;15(4):575–91.

8. Pallmann P, Bedding AW, Choodari-Oskooei B, Dimairo M, Flight L, Hampson LV, et al. Adaptive designs in clinical trials: why use them, and how to run and report them. BMC Med [Internet]. 2018 Dec [cited 2020 Apr 23];16(1). Available from: https://bmcmedicine.biomedcentral.com/articles/10.1186/s12916-018-1017-7

9. Research C for DE and. Adaptive Design Clinical Trials for Drugs and Biologics [Internet]. U.S. Food and Drug Administration. 2019 [cited 2019 Nov 18]. Available from: http://www.fda.gov/regulatory-information/search-fda-guidance-documents/adaptive-design-clinical-trials-drugs-and-biologics

10. Novak JE, Inrig JK, Patel UD, Califf RM, Szczech LA. Negative trials in nephrology: what can we learn? Kidney Int. 2008 Nov;74(9):1121–7.

11. Chaitman BR. Effects of Ranolazine With Atenolol, Amlodipine, or Diltiazem on Exercise Tolerance and Angina Frequency in Patients With Severe Chronic AnginaA Randomized Controlled Trial. JAMA. 2004 Jan 21;291(3):309.

12. Wanner C, Krane V, März W, Olschewski M, Mann JFE, Ruf G, et al. Atorvastatin in Patients with Type 2 Diabetes Mellitus Undergoing Hemodialysis. N Engl J Med. 2005 Jul 21;353(3):238–48.

13. Cheung BMY, Lauder IJ, Lau C-P, Kumana CR. Meta-analysis of large randomized controlled trials to evaluate the impact of statins on cardiovascular outcomes. Br J Clin Pharmacol. 2004 May;57(5):640–51.

14. Velenosi TJ, Urquhart BL. Pharmacokinetic considerations in chronic kidney disease and patients requiring dialysis. Expert Opin Drug Metab Toxicol. 2014 Aug;10(8):1131–43.

15. Pushpakom S, Kolamunnage-Dona R, Taylor C, Foster T, Spowart C, García-Fiñana M, et al. TAILoR (TelmisArtan and InsuLin Resistance in Human Immunodeficiency Virus [HIV]): An Adaptive-design, Dose-ranging Phase IIb Randomized Trial of Telmisartan for the Reduction of Insulin Resistance in HIV-positive Individuals on Combination Antiretroviral Therapy. Clin Infect Dis [Internet]. 2019 Jul 3 [cited 2020 Apr 23]; Available from: https://academic.oup.com/cid/advance-article/doi/10.1093/cid/ciz589/5527878

16. Moher D, Liberati A, Tetzlaff J, Altman DG, for the PRISMA Group. Preferred reporting items for systematic reviews and meta-analyses: the PRISMA statement. BMJ. 2009 Jul 21;339(jul21 1):b2535–b2535.

17. Judge C, Murphy RP, Cormican S, Smyth A, O’Halloran M, O’Donnell M. Adaptive design methods in dialysis clinical trials: a systematic review protocol. BMJ Open. 2020 Aug 27;10(8):e036755.

18. Beaubien-Souligny W, Kontar L, Blum D, Bouchard J, Denault AY, Wald R. Meta-analysis of Randomized Controlled Trials Using Tool-Assisted Target Weight Adjustments in Chronic Dialysis Patients. Kidney Int Rep. 2019 Jul;S2468024919314044.

19. Bothwell LE, Avorn J, Khan NF, Kesselheim AS. Adaptive design clinical trials: a review of the literature and ClinicalTrials.gov. BMJ Open. 2018 Feb 1;8(2):e018320.

20. Hatfield I, Allison A, Flight L, Julious SA, Dimairo M. Adaptive designs undertaken in clinical research: a review of registered clinical trials. Trials [Internet]. 2016 Dec [cited 2020 Aug 21];17(1). Available from: http://www.trialsjournal.com/content/17/1/150

21. Higgins JP. Revised Cochrane risk-of-bias tool for randomized trials (RoB 2). In RoB2 Development Group; 2019. Available from: https://sites.google.com/site/riskofbiastool/welcome/rob-2-0-tool/current-version-of-rob-2

22. Higgins JPT, Altman DG, Gotzsche PC, Juni P, Moher D, Oxman AD, et al. The Cochrane Collaboration’s tool for assessing risk of bias in randomised trials. BMJ. 2011 Oct 18;343(oct18 2):d5928–d5928.

23. Mistry P, Dunn JA, Marshall A. A literature review of applied adaptive design methodology within the field of oncology in randomised controlled trials and a proposed extension to the CONSORT guidelines. BMC Med Res Methodol. 2017 Jul 18;17(1):108.

24. Clayton JA, Arnegard ME. Taking cardiology clinical trials to the next level: A call to action. Clin Cardiol. 2018 Feb;41(2):179–84.

25. van Werkhoven CH, Harbarth S, Bonten MJM. Adaptive designs in clinical trials in critically ill patients: principles, advantages and pitfalls. Intensive Care Med. 2019 May 1;45(5):678–82.

26. Talisa VB, Yende S, Seymour CW, Angus DC. Arguing for Adaptive Clinical Trials in Sepsis. Front Immunol [Internet]. 2018 Jun 28 [cited 2021 Jan 14];9. Available from: https://www.ncbi.nlm.nih.gov/pmc/articles/PMC6031704/

27. Perkovic V, Craig JC, Chailimpamontree W, Fox CS, Garcia-Garcia G, Benghanem Gharbi M, et al. Action plan for optimizing the design of clinical trials in chronic kidney disease. Kidney Int Suppl. 2017 Oct;7(2):138–44.

28. Zoccali C, Vanholder R, Wagner CA, Anders H-J, Blankestijn PJ, Bruchfeld A, et al. Funding kidney research as a public health priority: challenges and opportunities. Nephrol Dial Transplant [Internet]. 2020 Sep 4 [cited 2021 Jan 14];(gfaa163). Available from: https://doi.org/10.1093/ndt/gfaa163

29. Bove T, Zangrillo A, Guarracino F, Alvaro G, Persi B, Maglioni E, et al. Effect of Fenoldopam on Use of Renal Replacement Therapy Among Patients With Acute Kidney Injury After Cardiac Surgery: A Randomized Clinical Trial. JAMA. 2014 Dec 3;312(21):2244.

30. Joannes-Boyau O, Honoré PM, Perez P, Bagshaw SM, Grand H, Canivet J-L, et al. High-volume versus standard-volume haemofiltration for septic shock patients with acute kidney injury (IVOIRE study): a multicentre randomized controlled trial. Intensive Care Med. 2013 Sep;39(9):1535–46.

31. Thiele H, Akin I, Sandri M, de Waha-Thiele S, Meyer-Saraei R, Fuernau G, et al. One-Year Outcomes after PCI Strategies in Cardiogenic Shock. N Engl J Med. 2018 Nov;379(18):1699–710.

32. Thiele H, Desch S, Piek JJ, Stepinska J, Oldroyd K, Serpytis P, et al. Multivessel versus culprit lesion only percutaneous revascularization plus potential staged revascularization in patients with acute myocardial infarction complicated by cardiogenic shock: Design and rationale of CULPRIT-SHOCK trial. Am Heart J. 2016 Feb;172:160–9.

33. Mehta RH, Leimberger JD, van Diepen S, Meza J, Wang A, Jankowich R, et al. Levosimendan in Patients with Left Ventricular Dysfunction Undergoing Cardiac Surgery. N Engl J Med. 2017 May 25;376(21):2032–42.

34. Mehta RH, Van Diepen S, Meza J, Bokesch P, Leimberger JD, Tourt-Uhlig S, et al. Levosimendan in patients with left ventricular systolic dysfunction undergoing cardiac surgery on cardiopulmonary bypass: Rationale and study design of the Levosimendan in Patients with Left Ventricular Systolic Dysfunction Undergoing Cardiac Surgery Requiring Cardiopulmonary Bypass (LEVO-CTS) trial. Am Heart J. 2016 Dec;182:62–71.

35. Intensity of Renal Support in Critically Ill Patients with Acute Kidney Injury. N Engl J Med. 2008 Jul 3;359(1):7–20.

36. Sharma S, Kelly YP, Palevsky PM, Waikar SS. Intensity of Renal Replacement Therapy and Duration of Mechanical Ventilation. Chest. 2020 Oct;158(4):1473–81.

37. Ejaz AA, Martin TD, Johnson RJ, Winterstein AG, Klodell CT, Hess PJ, et al. Prophylactic nesiritide does not prevent dialysis or all-cause mortality in patients undergoing high-risk cardiac surgery. J Thorac Cardiovasc Surg. 2009 Oct;138(4):959–64.

38. Acker CG, Singh AR, Flick RP, Bernardini J, Greenberg A, Johnson JP. A trial of thyroxine in acute renal failure. Kidney Int. 2000 Jan;57(1):293–8.

39. Gaudry S, Hajage D, Schortgen F, Martin-Lefevre L, Pons B, Boulet E, et al. Initiation Strategies for Renal-Replacement Therapy in the Intensive Care Unit. N Engl J Med. 2016 Jul 14;375(2):122–33.

40. Gaudry S, Hajage D, Schortgen F, Martin-Lefevre L, Tubach F, Pons B, et al. Comparison of two strategies for initiating renal replacement therapy in the intensive care unit: study protocol for a randomized controlled trial (AKIKI). Trials [Internet]. 2015 Dec [cited 2020 Oct 27];16(1). Available from: http://trialsjournal.biomedcentral.com/articles/10.1186/s13063-015-0718-x

41. Weisbord SD, Gallagher M, Jneid H, Garcia S, Cass A, Thwin S-S, et al. Outcomes after Angiography with Sodium Bicarbonate and Acetylcysteine. N Engl J Med. 2018 Feb 15;378(7):603–14.

42. Schanz M, Wasser C, Allgaeuer S, Schricker S, Dippon J, Alscher MD, et al. Urinary [TIMP-2]·[IGFBP7]-guided randomized controlled intervention trial to prevent acute kidney injury in the emergency department. Nephrol Dial Transplant. 2019 Nov 1;34(11):1902–9.

43. Zarbock A, Küllmar M, Kindgen-Milles D, Wempe C, Gerss J, Brandenburger T, et al. Effect of Regional Citrate Anticoagulation vs Systemic Heparin Anticoagulation During Continuous Kidney Replacement Therapy on Dialysis Filter Life Span and Mortality Among Critically Ill Patients With Acute Kidney Injury: A Randomized Clinical Trial. JAMA. 2020 Oct 27;324(16):1629.

44. Meersch M, Küllmar M, Wempe C, Kindgen-Milles D, Kluge S, Slowinski T, et al. Regional citrate versus systemic heparin anticoagulation for continuous renal replacement therapy in critically ill patients with acute kidney injury (RICH) trial: study protocol for a multicentre, randomised controlled trial. BMJ Open. 2019 Jan;9(1):e024411.

45. Combes A, Bréchot N, Amour J, Cozic N, Lebreton G, Guidon C, et al. Early High-Volume Hemofiltration versus Standard Care for Post–Cardiac Surgery Shock. The HEROICS Study. Am J Respir Crit Care Med. 2015 Nov 15;192(10):1179–90.

46. Robinson S, Zincuk A, Larsen UL, Ekstrøm C, Toft P. A feasible strategy for preventing blood clots in critically ill patients with acute kidney injury (FBI): study protocol for a randomized controlled trial. Trials [Internet]. 2014 Dec [cited 2020 Oct 27];15(1). Available from: https://trialsjournal.biomedcentral.com/articles/10.1186/1745-6215-15-226

47. Zarbock A, Kellum JA, Schmidt C, Van Aken H, Wempe C, Pavenstädt H, et al. Effect of Early vs Delayed Initiation of Renal Replacement Therapy on Mortality in Critically Ill Patients With Acute Kidney Injury: The ELAIN Randomized Clinical Trial. JAMA. 2016 May 24;315(20):2190.

48. Zarbock A, Gerß J, Van Aken H, Boanta A, Kellum JA, Meersch M. Early versus late initiation of renal replacement therapy in critically ill patients with acute kidney injury (The ELAIN-Trial): study protocol for a randomized controlled trial. Trials [Internet]. 2016 Dec [cited 2020 Oct 27];17(1). Available from: http://www.trialsjournal.com/content/17/1/148

49. Dember LM, Beck GJ, Allon M, Delmez JA, Dixon BS, Greenberg A, et al. Effect of Clopidogrel on Early Failure of Arteriovenous Fistulas for Hemodialysis: A Randomized Controlled Trial. JAMA. 2008 May 14;299(18):2164.

50. Irish AB, Viecelli AK, Hawley CM, Hooi L-S, Pascoe EM, Paul-Brent P-A, et al. Effect of Fish Oil Supplementation and Aspirin Use on Arteriovenous Fistula Failure in Patients Requiring Hemodialysis: A Randomized Clinical Trial. JAMA Intern Med. 2017 Feb 1;177(2):184.

51. Viecelli AK, Polkinghorne KR, Pascoe EM, Paul-Brent P-A, Hawley CM, Badve SV, et al. Fish oil and aspirin effects on arteriovenous fistula function: Secondary outcomes of the randomised omega-3 fatty acids (Fish oils) and Aspirin in Vascular access OUtcomes in REnal Disease (FAVOURED) trial. Puebla I, editor. PLOS ONE. 2019 Mar 26;14(3):e0213274.

52. Chapman WC, Singla N, Genyk Y, McNeil JW, Renkens KL, Reynolds TC, et al. A Phase 3, Randomized, Double-Blind Comparative Study of the Efficacy and Safety of Topical Recombinant Human Thrombin and Bovine Thrombin in Surgical Hemostasis. J Am Coll Surg. 2007 Aug;205(2):256–65.

53. Ismail-Beigi F, Craven T, Banerji MA, Basile J, Calles J, Cohen RM, et al. Effect of intensive treatment of hyperglycaemia on microvascular outcomes in type 2 diabetes: an analysis of the ACCORD randomised trial. The Lancet. 2010 Aug;376(9739):419–30.

54. Freedman BI, Wuerth J-P, Cartwright K, Bain RP, Dippe S, Hershon K, et al. Design and Baseline Characteristics for the Aminoguanidine Clinical Trial in Overt Type 2 Diabetic Nephropathy (ACTION II). Control Clin Trials. 1999 Oct;20(5):493–510.

55. Johnson DW, Badve SV, Pascoe EM, Beller E, Cass A, Clark C, et al. Antibacterial honey for the prevention of peritoneal-dialysis-related infections (HONEYPOT): a randomised trial. Lancet Infect Dis. 2014 Jan;14(1):23–30.

56. Pascoe EM, Lo S, Scaria A, Badve SV, Beller EM, Cass A, et al. The Honeypot Randomized Controlled Trial Statistical Analysis Plan. Perit Dial Int J Int Soc Perit Dial. 2013 Jul;33(4):426–35.

57. Besarab A, Bolton WK, Browne JK, Egrie JC, Nissenson AR, Okamoto DM, et al. The Effects of Normal as Compared with Low Hematocrit Values in Patients with Cardiac Disease Who Are Receiving Hemodialysis and Epoetin. N Engl J Med. 1998 Aug 27;339(9):584–90.

58. Dixon BS, Beck GJ, Vazquez MA, Greenberg A, Delmez JA, Allon M, et al. Effect of Dipyridamole plus Aspirin on Hemodialysis Graft Patency. N Engl J Med. 2009 May 21;360(21):2191–201.

59. Torres VE, Abebe KZ, Chapman AB, Schrier RW, Braun WE, Steinman TI, et al. Angiotensin Blockade in Late Autosomal Dominant Polycystic Kidney Disease. N Engl J Med. 2014 Dec 11;371(24):2267–76.

60. Perkovic V, Jardine MJ, Neal B, Bompoint S, Heerspink HJL, Charytan DM, et al. Canagliflozin and Renal Outcomes in Type 2 Diabetes and Nephropathy. N Engl J Med. 2019 Jun 13;380(24):2295–306.

61. Wiviott SD, Raz I, Bonaca MP, Mosenzon O, Kato ET, Cahn A, et al. Dapagliflozin and Cardiovascular Outcomes in Type 2 Diabetes. N Engl J Med. 2019 Jan 24;380(4):347–57.

62. Knoll GA, Fergusson D, Chassé M, Hebert P, Wells G, Tibbles LA, et al. Ramipril versus placebo in kidney transplant patients with proteinuria: a multicentre, double-blind, randomised controlled trial. Lancet Diabetes Endocrinol. 2016 Apr;4(4):318–26.

63. Knoll GA, Cantarovitch M, Cole E, Gill J, Gourishankar S, Holland D, et al. The Canadian ACE-inhibitor trial to improve renal outcomes and patient survival in kidney transplantation study design. Nephrol Dial Transplant. 2007 Aug 17;23(1):354–8.

64. Blankestijn PJ, Fischer KI, Barth C, Cromm K, Canaud B, Davenport A, et al. Benefits and harms of high-dose haemodiafiltration versus high-flux haemodialysis: the comparison of high-dose haemodiafiltration with high-flux haemodialysis (CONVINCE) trial protocol. BMJ Open. 2020 Feb;10(2):e033228.

65. Fellström BC, Jardine AG, Schmieder RE, Holdaas H, Bannister K, Beutler J, et al. Rosuvastatin and Cardiovascular Events in Patients Undergoing Hemodialysis. N Engl J Med. 2009 Apr 2;360(14):1395–407.

66. Fellström B, Holdaas H, Jardine AG, Rose H, Schmieder R, Wilpshaar W, et al. Effect of Rosuvastatin on Outcomes in Chronic Haemodialysis Patients: Baseline Data from the AURORA Study. Kidney Blood Press Res. 2007;30(5):314–22.

67. Grooteman MPC, van den Dorpel MA, Bots ML, Penne EL, van der Weerd NC, Mazairac AHA, et al. Effect of Online Hemodiafiltration on All-Cause Mortality and Cardiovascular Outcomes. J Am Soc Nephrol. 2012 Jun;23(6):1087–96.

68. the CONTRAST study group, Penne EL, Blankestijn PJ, Bots ML, van den Dorpel MA, Grooteman MP, et al. Effect of increased convective clearance by on-line hemodiafiltration on all cause and cardiovascular mortality in chronic hemodialysis patients – the Dutch CONvective TRAnsport STudy (CONTRAST): rationale and design of a randomised controlled trial [ISRCTN38365125]. Curr Control Trials Cardiovasc Med [Internet]. 2005 Dec [cited 2020 Oct 27];6(1). Available from: http://trialsjournal.biomedcentral.com/articles/10.1186/1468-6708-6-8

69. Karunanithy N, Mesa IR, Dorling A, Calder F, Katsanos K, Semik V, et al. Paclitaxel-coated balloon fistuloplasty versus plain balloon fistuloplasty only to preserve the patency of arteriovenous fistulae used for haemodialysis (PAVE): study protocol for a randomised controlled trial. Trials [Internet]. 2016 Dec [cited 2020 Oct 27];17(1). Available from: http://trialsjournal.biomedcentral.com/articles/10.1186/s13063-016-1372-7

70. Kopple JD, Cheung AK, Christiansen JS, Djurhuus CB, El Nahas M, Feldt-Rasmussen B, et al. OPPORTUNITYC: a large-scale randomized clinical trial of growth hormone in hemodialysis patients. Nephrol Dial Transplant. 2011 Dec 1;26(12):4095–103.

71. Kopple JD, Cheung AK, Christiansen JS, Djurhuus CB, El Nahas M, Feldt-Rasmussen B, et al. OPPORTUNITY ™: A Randomized Clinical Trial of Growth Hormone on Outcome in Hemodialysis Patients. Clin J Am Soc Nephrol. 2008 Nov;3(6):1741–51.

72. Kwiatkowski DM, Goldstein SL, Cooper DS, Nelson DP, Morales DLS, Krawczeski CD. Peritoneal Dialysis vs Furosemide for Prevention of Fluid Overload in Infants After Cardiac Surgery: A Randomized Clinical Trial. JAMA Pediatr. 2017 Apr 1;171(4):357.

73. Lemkes JS, Janssens GN, van der Hoeven NW, Jewbali LSD, Dubois EA, Meuwissen M, et al. Coronary Angiography after Cardiac Arrest without ST-Segment Elevation. N Engl J Med. 2019 Apr 11;380(15):1397–407.

74. Lemkes JS, Janssens GN, Straaten HMO, Elbers PW, van der Hoeven NW, Tijssen JGP, et al. Coronary angiography after cardiac arrest: Rationale and design of the COACT trial. Am Heart J. 2016 Oct;180:39–45.

75. Douglas IS, Alapat PM, Corl KA, Exline MC, Forni LG, Holder AL, et al. Fluid Response Evaluation in Sepsis Hypotension and Shock. Chest. 2020 Oct;158(4):1431–45.

76. Vinsonneau C, Camus C, Combes A, Costa de Beauregard MA, Klouche K, Boulain T, et al. Continuous venovenous haemodiafiltration versus intermittent haemodialysis for acute renal failure in patients with multiple-organ dysfunction syndrome: a multicentre randomised trial. The Lancet. 2006 Jul;368(9533):379–85.

77. Pettilä V, Merz T, Wilkman E, Perner A, Karlsson S, Lange T, et al. Targeted tissue perfusion versus macrocirculation-guided standard care in patients with septic shock (TARTARE-2S): study protocol and statistical analysis plan for a randomized controlled trial. Trials [Internet]. 2016 Dec [cited 2020 Oct 27];17(1). Available from: http://trialsjournal.biomedcentral.com/articles/10.1186/s13063-016-1515-x

78. The ANDROMEDA-SHOCK Study Investigators, Hernández G, Cavalcanti AB, Ospina-Tascón G, Zampieri FG, Dubin A, et al. Early goal-directed therapy using a physiological holistic view: the ANDROMEDA-SHOCK—a randomized controlled trial. Ann Intensive Care [Internet]. 2018 Dec [cited 2020 Oct 27];8(1). Available from: https://annalsofintensivecare.springeropen.com/articles/10.1186/s13613-018-0398-2

79. Tumlin JA, Galphin CM, Tolwani AJ, Chan MR, Vijayan A, Finkel K, et al. A Multi-Center, Randomized, Controlled, Pivotal Study to Assess the Safety and Efficacy of a Selective Cytopheretic Device in Patients with Acute Kidney Injury. Cravedi P, editor. PLOS ONE. 2015 Aug 5;10(8):e0132482.

80. Riley AA, Jefferies JL, Nelson DP, Bennett MR, Blinder JJ, Ma Q, et al. Peritoneal Dialysis does not Adversely Affect Kidney Function Recovery after Congenital Heart Surgery. Int J Artif Organs. 2014 Jan;37(1):39–47.

81. Hayashi T, Maruyama S, Nangaku M, Narita I, Hirakata H, Tanabe K, et al. Darbepoetin Alfa in Patients with Advanced CKD without Diabetes: Randomized, Controlled Trial. Clin J Am Soc Nephrol. 2020 May 7;15(5):608–15.

82. Imai E, Maruyama S, Nangaku M, Hirakata H, Hayashi T, Narita I, et al. Rationale and study design of a randomized controlled trial to assess the effects of maintaining hemoglobin levels using darbepoetin alfa on prevention of development of end-stage kidney disease in non-diabetic CKD patients (PREDICT Trial). Clin Exp Nephrol. 2016 Feb;20(1):71–6.

83. Marsen TA, Beer J, Mann H. Intradialytic parenteral nutrition in maintenance hemodialysis patients suffering from protein-energy wasting. Results of a multicenter, open, prospective, randomized trial. Clin Nutr. 2017 Feb;36(1):107–17.

84. Fishbane S, Jamal A, Munera C, Wen W, Menzaghi F. A Phase 3 Trial of Difelikefalin in Hemodialysis Patients with Pruritus. N Engl J Med. 2020 Jan 16;382(3):222–32.

85. Kammerer T, Brettner F, Hilferink S, Hulde N, Klug F, Pagel J-I, et al. No Differences in Renal Function between Balanced 6% Hydroxyethyl Starch (130/0.4) and 5% Albumin for Volume Replacement Therapy in Patients Undergoing Cystectomy. Anesthesiology. 2018 Jan 1;128(1):67–78.

86. Kammerer T, Klug F, Schwarz M, Hilferink S, Zwissler B, von Dossow V, et al. Comparison of 6 % hydroxyethyl starch and 5 % albumin for volume replacement therapy in patients undergoing cystectomy (CHART): study protocol for a randomized controlled trial. Trials [Internet]. 2015 Dec [cited 2020 Oct 27];16(1). Available from: http://trialsjournal.biomedcentral.com/articles/10.1186/s13063-015-0866-z

87. Kratochwill K, Boehm M, Herzog R, Gruber K, Lichtenauer AM, Kuster L, et al. Addition of Alanyl-Glutamine to Dialysis Fluid Restores Peritoneal Cellular Stress Responses – A First-In-Man Trial. Eller K, editor. PLOS ONE. 2016 Oct 21;11(10):e0165045.

88. Fujimoto K, Adachi H, Yamazaki K, Nomura K, Saito A, Matsumoto Y, et al. Comparison of the pain-reducing effects of EMLA cream and of lidocaine tape during arteriovenous fistula puncture in patients undergoing hemodialysis: A multi-center, open-label, randomized crossover trial. Wu P-H, editor. PLOS ONE. 2020 Mar 25;15(3):e0230372.

89. Himmelfarb J, Chertow GM, McCullough PA, Mesana T, Shaw AD, Sundt TM, et al. Perioperative THR-184 and AKI after Cardiac Surgery. J Am Soc Nephrol. 2018 Feb;29(2):670–9.

90. Hosgood SA, Saeb-Parsy K, Wilson C, Callaghan C, Collett D, Nicholson ML. Protocol of a randomised controlled, open-label trial of ex vivo normothermic perfusion versus static cold storage in donation after circulatory death renal transplantation. BMJ Open. 2017 Jan;7(1):e012237.

91. Bestle MH, Clausen NE, Søe-Jensen P, Kristiansen KT, Lange T, Johansson PI, et al. Efficacy and safety of iloprost in patients with septic shock-induced endotheliopathy—Protocol for the multicenter randomized, placebo-controlled, blinded, investigator-initiated trial. Acta Anaesthesiol Scand. 2020 May;64(5):705–11.

92. Pickkers P, Mehta RL, Murray PT, Joannidis M, Molitoris BA, Kellum JA, et al. Effect of Human Recombinant Alkaline Phosphatase on 7-Day Creatinine Clearance in Patients With Sepsis-Associated Acute Kidney Injury: A Randomized Clinical Trial. JAMA. 2018 Nov 20;320(19):1998.

93. E P, RL M, PT M, J H, M J, JA K, et al. Study protocol for a multicentre randomised controlled trial: Safety, Tolerability, efficacy and quality of life Of a human recombinant alkaline Phosphatase in patients with sepsis-associated Acute Kidney Injury (STOP-AKI). BMJ Open. 2016;6(9):e012371.

94. Kassimatis T, Qasem A, Douiri A, Ryan EG, Rebollo-Mesa I, Nichols LL, et al. A double-blind randomised controlled investigation into the efficacy of Mirococept (APT070) for preventing ischaemia reperfusion injury in the kidney allograft (EMPIRIKAL): study protocol for a randomised controlled trial. Trials [Internet]. 2017 Dec [cited 2020 Oct 27];18(1). Available from: http://trialsjournal.biomedcentral.com/articles/10.1186/s13063-017-1972-x

