## Supplementary Appendix for "Adaptive design methods in dialysis clinical trials – a systematic review"

### Supplementary Table 1 – Search strategy for Medline (Pubmed)

dialysis[tiab]

OR

peritoneal dialysis[tiab]

OR

hemodialysis[tiab]

OR

hemodiafiltration[tiab]

OR

haemodiafiltration[tiab]

OR

hemofiltration[tiab]

OR

haemofiltration

OR

extracorporeal blood cleansing[tiab]

OR

haemodialysis[tiab]

OR

Renal Dialysis[mh]

OR

Renal replacement[tiab]

OR

end stage kidney[tiab]

OR

end stage renal[tiab]

OR

stage 5 kidney[tiab]

OR

stage 5 renal[tiab]

### Supplementary Table 2 – Search strategy for Recoll (Full text search)

phase ii/iii[tiab]

OR

treatment switching[tiab]

OR

biomarker adaptive[tiab]

OR

biomarker adaptive design[tiab]

OR

biomarker adjusted[tiab]

OR

adaptive hypothesis[tiab]

OR

adaptive dose-finding[tiab]

OR

pick-the winner[tiab]

OR

drop-the-loser[tiab]

OR

sample size re-estimation[tiab]

OR

re-estimations[tiab]

OR

adaptive randomization[tiab]

OR

group sequential[tiab]

OR

adaptive seamless[tiab]

OR

adaptive design[tiab]

OR

Interim monitoring[tiab]

OR

Bayesian adaptive[tiab]

OR

Flexible design[tiab]

OR

Adaptive trial[tiab]

OR

play-the-winner[tiab]

OR

adaptive method[tiab]

OR

(adaptive[All Fields] AND dose[All Fields] AND adjusting[All Fields])

OR

response adaptive[All Fields]

OR

adaptive allocation[All Fields]

OR

adaptive signature design[tiab]

OR

treatment adaptive[tiab]

OR

covariate adaptive[tiab]

OR

sample size adjustment[tiab].

### Supplementary Table 3 – Characteristics of the trials

| **Study Characteristic** | **Categories** | **Description** |
| --- | --- | --- |
| Nature of Adaptive Design | GSD/SSR/DS/DE/Seamless/Interim Analysis | The type of adaptive design used in the trial. |
| Stopping Rule | Futility/Efficacy/Two sided/ N/A | If a stopping rule was used, what was the nature of the stopping rule. |
| Year of study completion | None | The year of study completion. |
| Population under study | None | A description of the population studied e.g. patients with diabetes. |
| Chronicity of RRT | Acute Kidney Injury (AKI) / ESKD | A category for the chronicity of Renal Replacement Therapy (RRT), either Acute Kidney Injury (AKI) or End Stage Kidney Disease (ESKD). |
| Intervention | None | A free text description of the intervention. |
| Nature of the intervention | Medication/Medical Device/Dialysis Parameter | A category for the nature of the intervention. |
| Primary Outcome | None | A description of the primary outcome of the trial. |
| Type of primary outcome | Continuous or dichotomous | A categorial variable for the type of primary outcome variable. |
| Nature of primary outcome | Surrogate, patient-centred or hard clinical | A categorial variable for the nature of primary outcome variable either surrogate, patient-centred or hard clinical. |
| Dialysis Modality | Haemodialysis, peritoneal dialysis, haemodiafiltration or haemofiltration | A categorial variable for the dialysis modality. |
| Sample Size of Study | None | The number of participants in the study. |
| The country of the lead investigator. | None | The country of the lead investigator. |
| The funder of the study | Public/Private | A categorial variable for source of funding for the study. |
| Study Phase | Phase II/Phase III/Combined Phase II/III | A categorial variable for study phase. |

### Supplementary Table 4 – Risk of Bias Assessment

| **Experimental** | **Comparator** | **Randomization process** | **Deviations from intended interventions** | **Missing outcome data** | **Measurement of the outcome** | **Selection of the reported result** | **Overall Bias** |
| --- | --- | --- | --- | --- | --- | --- | --- |
| Fenoldopam infusion | Placebo (saline) | Low | Low | Low | Low | Low | Low |
| Clopidogrel | Placebo | Low | Low | Low | Low | Low | Low |
| Fish Oil Supplementation and Aspirin Use | Placebo | Low | Low | Low | Low | Low | Low |
| Peritoneal Dialysis | Furosemide | Low | Low | Low | Low | Low | Low |
| High-volume haemofiltration | Standard-volume haemofiltration | Some concerns | Some concerns | Low | Low | Low | Some concerns |
| Culprit-lesion-only PCI | Immediate multivessel PCI | Low | High | Low | Low | Low | High |
| Immediate coronary angiography | Delayed coronary angiography | Low | High | Low | Low | Low | High |
| Levosimendan | Placebo | Low | Low | Low | Low | Low | Low |
| Fluid Response Evaluation | Usual Care | Low | High | High | Low | Low | High |
| Intensive RRT | Less Intensive RRT | Low | High | Low | Low | Some concerns | High |
| Intradialytic parenteral nutrition | standardized nutritional counseling | Low | Low | Low | Low | High | High |
| Topical Recombinant Human Thrombin | Bovine Thrombin | Low | Low | Low | Low | Some concerns | Some concerns |
| Nesiritide | Placebo | Low | Some concerns | Low | Low | Some concerns | Some concerns |
| Continuous venovenous haemodiafiltration | Intermittent haemodialysis | Low | High | Low | Low | Some concerns | High |
| Intensive glycemic therapy with a target HbA1c of <6.0% | standard therapy with a target of 7-7.9% | Low | Low | Low | Low | Low | Low |
| Antibacterial honey | standard exit-site care | Low | High | Low | Some concerns | Low | High |
| Thyroxine | Placebo | Some concerns | Low | Low | Low | Some concerns | Some concerns |
| Normal Hematocrit Values | Low Hematocrit Values | Some concerns | Low | Some concerns | Low | Some concerns | Some concerns |
| Dipyridamole plus aspirin | Placebo | Low | Low | Low | Low | Low | Low |
| Angiotensin Blockade | Placebo | Low | Low | Low | Low | Low | High |
| Early phase RRT | Delayed phase RRT | Low | Low | Low | Low | Low | Low |
| Sodium bicarbonate | Normal saline and acetylcysteine | Low | Low | Low | Low | Low | Low |
| Canagliflozin | Placebo | Low | Low | Low | Low | Low | Low |
| Dapagliflozin | Placebo | Low | Low | Low | Low | Low | Low |
| difelikefalin | placebo | Low | Low | Low | Low | Low | Low |
| Ramipril | Placebo | Some concerns | Low | Low | Low | Low | Some concerns |
| Renal consult | Usual care | Low | Low | Low | Low | Low | Low |
| Hydroxyethyl starch | 5% albumin | Low | Some concerns | Some concerns | Low | Low | Some concerns |
| Recombinant alkaline phosphatase 0.4mg/kg | recombinant alkaline phosphatase 1.6mg/kg or placebo | Low | Low | Low | Low | Low | Low |
| Regional citrate anticoagulation | Systemic heparin | Low | Some concerns | Low | Low | Low | Some concerns |
| online hemodiafiltration | low flux haemofiltration | Low | Low | Low | Low | Low | Low |
| Early RRT | Delayed RRT | Some concerns | Low | Low | Low | Low | Some concerns |
| Selective cytopheretic device and CVVHD | CVVHD | Low | Low | High | Low | Low | High |
| alagn | glucose based pdf | Low | Low | Low | Low | Low | Low |
| THR 184 | placebo | Some concerns | Low | Low | Low | Low | Some concerns |
| Recombinant hGH | placebo | Some concerns | Low | Low | Low | Low | Some concerns |
| high-hemoglobin | low-hemoglobin | Low | Low | Low | High | Low | High |
| Continue CPD | Discontinue CPD | Some concerns | Low | Low | Some concerns | Low | Some concerns |
| rosuvastatin | placebo | Some concerns | Low | Low | Low | Low | Some concerns |
| Early high volume haemofiltration | Standard care | Some concerns | Low | Low | Low | Low | Low |

### Supplementary Figure 1 – Risk of Bias Assessment of Dialysis Randomized Clinical Trials with Adaptive Designs

As percentage (intention-to-treat)

|  | Low risk | Some concerns | High risk |
| --- | --- | --- | --- |
| Randomization process | 75 | 25 | 0 |
| Deviations from intended interventions | 75 | 10 | 15 |
| Mising outcome data | 90 | 5 | 5 |
| Measurement of the outcome | 92.5 | 5 | 2.5 |
| Selection of the reported result | 82.5 | 15 | 2.5 |
| Overall Bias | 42.5 | 32.5 | 25 |
